# Reduced Expression of MTSS1 Increases Sarcomere Number and Improves Contractility in Select Forms of Monogenic DCM

**DOI:** 10.1101/2024.08.14.24311020

**Authors:** Hannah Kleppe, Anastasia Budan, Luke Zhang, Marie Majetic, Reva Shenwai, Alan Levinson, Olga Cisne-Thompson, Farshad Farshidfar, Jonathan Tsui, Sylwia Figarska, Tim Hoey, James Priest, Rebecca Slater

## Abstract

**Background:** The I-bar protein MTSS1 is a known modifier of heart failure and contractile phenotypes but its role in modulating contractile dysfunction in genetic forms of Mendelian dilated cardiomyopathy (DCM) is not known.

**Methods:** The potential role of cardiac MTSS1 in TTN DCM was explored using time-to-event models in longitudinal human datasets. Using induced siRNA and mutant forms of pluripotent stem cell cardiomyocytes (iPSC-CMs) the impact of siRNA knockdown of MTSS upon sarcomere and Cardiomyocyte biology was assessed via quantitative high-content microscopy, and the impact and mechanism of MTSS1 knockdown upon contractility was assessed using engineered heart tissues (EHTs).

**Results:** Amongst individuals affected with TTN DCM, a variant conferring lower cardiac levels of MTSS1 was associated with significantly improved event-free survival from cardiovascular death or heart transplant (HR 0.29, p=0.0016). Knockdown of MTSS1 by siRNA significantly improved the appearance of iPSC-CM models of TTN (p=2.9e-06), CSRP3 (p=3.1e-14), and RBM20 (p=4.4e-04) DCM as assessed by quantitative microscopy. Correspondingly, siRNA knockdown of MTSS1 increased contractility in EHT models of TTN DCM (p=0.003), CSRP3 DCM (p=0.008), and RBM20 DCM (p<2e-16). Across all genetic backgrounds, knockdown of MTSS1 was observed to increase the number of sarcomeres (p<0.0001), and in co-immunoprecipitation experiments MTSS1 physically interacts with MYO18A a key determinant of early sarcomere formation. Knockdown of MTSS1 resulted in increased transcription of MYH7 (0.29 log_2_FC, p=2.9e-06) along with other sarcomere genes.

**Conclusions:** In iPSC-CMs Knockdown of MTSS1 by siRNA increased number of sarcomeres and was observed to increase twitch force in select in vitro models, and may suggest MTSS1 plays a previously unrecognized role in modulating sarcomere production. Human observational and iPSC-CM experimental data supports the hypothesis that reduced expression of MTSS1 may be beneficial in Mendelian DCM caused by TTN, RBM20, and CSRP3.

## INTRODUCTION

Dilated Cardiomyopathy (DCM) is a form of heart failure typically characterized by reduced cardiac output and thinning of ventricular chambers affecting up to 1 in 250 adults ^1^. A heritable cause of DCM is present in 50% of individuals, and Titin (TTN) is the most common DCM locus with truncating mutations found in approximately 25% of familial DCM and 18% of sporadic DCM cases^2^, followed by mutations in a variety of sarcomere and non-sarcomere genes such as BAG3, TPM1 and CSRP3 which comprise less common genetic forms of DCM^3^. There are no approved therapies which directly address the decreased contractility and sarcomere dysfunction which underlies DCM.

Genome wide association studies (GWAS) have identified MTSS1 as a modifier of cardiac function whereby genetic variation in a nearby regulatory region appears to lower cardiac expression of MTSS1 and is strongly and reproducibly associated with increased contractility and decreased risk for heart failure^4–8^. The I-BAR protein MTSS1 was first identified as a metastasis suppressor missing in metastatic bladder carcinoma cell lines and is known to have several functions including a role in cell motility along with actin assembly and integrity^9,10^. A mouse knockout of Mtss1 shows decreased LV end-diastolic dimensions and LV end-systolic dimension as well as trends towards increased LV fractional shortening^4^ and appeared to have a modest beneficial effect upon contractility when crossed with a Tpm1 D230N mouse model of DCM^11^.

Here, we examined the relationship of a human genetic modifier of cardiovascular function and disease risk MTSS1, to uncover a new potential treatment for specific genetic forms of dilated cardiomyopathy (DCM). We demonstrate the specific effect of naturally occurring genetic variation upon MTSS1 expression in a group of individuals affected with TTN DCM, and demonstrate improvements in contractility after MTSS1 knockdown in induced pluripotent stem cell (iPSC) models DCM. Furthermore, we additionally characterized the impact of MTSS1 upon the subcellular architecture of iPSC cardiomyocytes and propose a previously unrecognized role for MTSS1 modulating cardiomyocyte biology.

## RESULTS

### Human Genetics

The genomic mechanism for relationship of MTSS1 to cardiac phenotypes is hypothesized to be a disruption of a cardiac enhancer by a common genetic variant, confirmed by expressed quantitative trait (eQTL) analyses of multiple datasets where the rs35006907-C allele was associated with higher risk of DCM and increased MTSS1 expression in European and Han Chinese populations^4,12^. Data from the Genotype Tissue Expression Consortium confirms this finding where the T allele of rs12541595 is strongly associated with lower left ventricular expression of MTSS1 [Fig.1A] and is in strong linkage with rs35006907 (R2 0.9799). Within the UK Biobank, a large prospective cohort of participants enrolled at age 40 or later, 99 individuals carrying a pathogenic or likely pathogenic (P/LP) mutation in TTN developed a diagnosis of DCM^13^. Individuals carrying both TTN P/LP variants and the MTSS1 expression-lowering T allele of rs12541595, showed significantly improved event-free survival from cardiovascular death or heart transplant (HR 0.29, p=0.0016) relative to individuals homozygous for the G allele [Fig.1B] suggesting that lower levels of MTSS1 may also be specifically protective from the contractile dysfunction which underlies TTN DCM.

**Figure 1.**
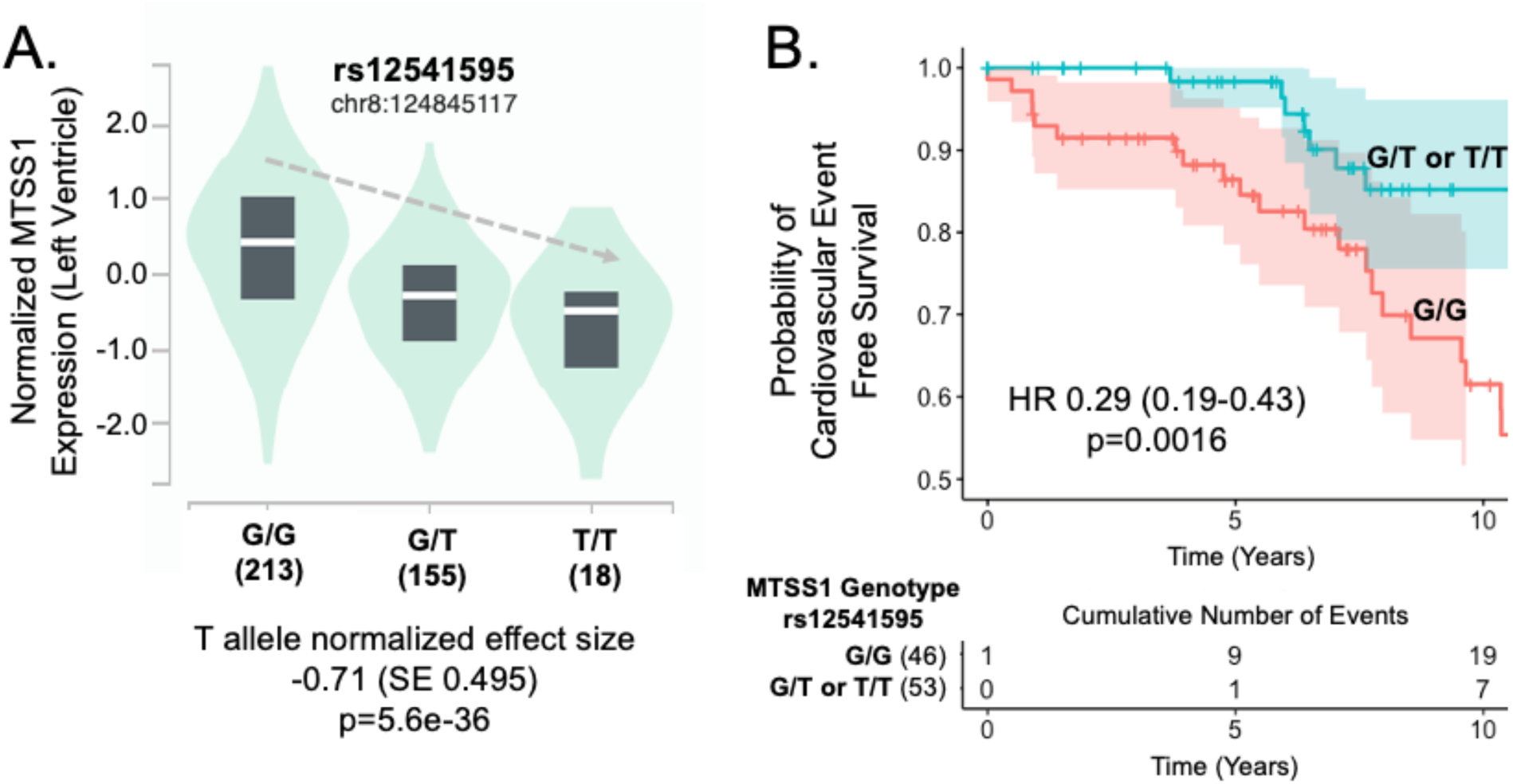
Lower cardiac expression of MTSS1 is associated with improved survival in TTN DCM. **A.** The T allele of rs12541595 is strongly associated with lower expression of MTSS1 in left ventricular cardiac tissue and to a lesser extent within atrial tissue (−0.28 normalized expression per T allele, p=9e-08). Data adapted from GTeX v8 database (accessed 2/5/24). **B.** Among individuals in the UK Biobank with TTN P/LP variants who developed DCM, the presence of an MTSS1 expression-lowering variant rs12541595 was observed to confer significantly improved event-free survival from cardiovascular death or heart transplant (HR 0.29, p=0.0016) in a model adjusted for age of DCM diagnosis and genetic sex.

### iPSC Models of DCM

To explore the impact of MTSS1 knockdown on cardiomyocyte biology we performed high-throughput high-content confocal microscopy to quantify aspects of cardiomyocyte and sarcomere biology (number of sarcomeres, sarcomere length, sarcomere angle, and sarcomere score/quality) based on previously described modifications of deep learning approach^14–16^. Using the imaging derived parameters, we developed a set of classification algorithms to incorporate the entirety of the sarcomere imaging data to distinguish untreated WT iPSC-CMs from DCM models of disease induced by siRNA knockdown. For each DCM model, a random forest classifier was constructed which incorporated and weighted the sarcomere biology data to output the probability that an image was derived from WT cells (treated with a scrambled siRNA) or a DCM disease gene [Fig 2A]. The prediction models displayed good or excellent discriminatory performance when evaluated on a held-out test dataset separate from the training data [Fig S1]. We observed that siRNA knockdown of MTSS1 shifted the classification of cells treated with siRNA-TTN (p=2.9e-06) and in a dose dependent effect for siRNA-CSRP3 (p_LM_dose_=1.34e-14) along with a dose dependent effect for RBM20 (p=5.7e-04) [Fig.2B,C,E]. By contrast, treatment with siRNA-MTSS1 only modestly shifted the model classifications of the WT iPSCs treated with siRNA for TPM1 with a much smaller dose-dependent effect (p=0.07) [Fig 2D]. Importantly, we observed that the siRNA models of DCM mildly impact iPSC-CM survival [Fig S2], therefore to ensure that the shift in classification was not solely attributable to an impact on cellular survival, siRNA MTSS1 was applied to an iPSC-CM line carrying a pathogenic DCM truncating mutation in TTN P22353X^17^ which also showed a moderate beneficial shift in classification when the siTTN-model was applied to the dataset (p=0.029) [Fig.S3].

**Figure 2.**
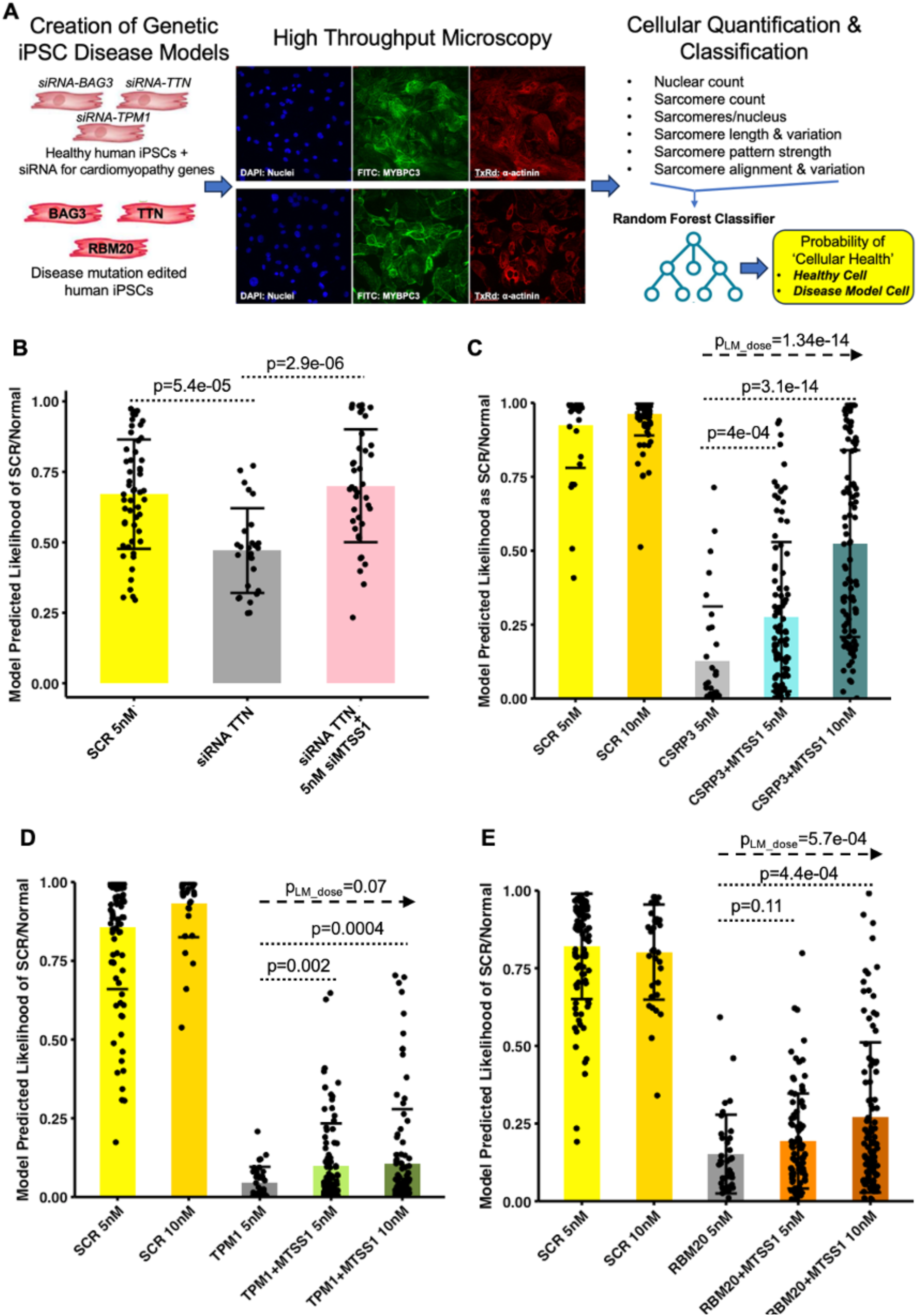
siRNA Knockdown of MTSS1 Improves the Appearance of iPSC Cardiomyocytes in Specific siRNA Models of Monogenic DCM. **A.** Diagram of experimental workflow where iPSC-CM models of monogenic DCM are treated with an siRNA for MTSS1 followed by imaging with three separate stains (DAPI, MYBPC3, and alpha-actinin). Five images per well are analyzed by a Matlab script to yield quantitative estimates of cellular parameters followed by usage for training a random forest algorithm which can output the probability that a selected image is derived from a normal cell or a model of DCM. **B.** 5nM siRNA-MTSS1 improves the model predicted appearance in an siRNA TTN model of DCM. **C.** siRNA-MTSS1 improves the model predicted appearance in an siRNA CSRP3 model of DCM in a dose-dependent manner. **D.** siRNA-MTSS1 does not meaningfully improve the model predicted appearance in an siRNA TPM1 model of DCM **E.** In an siRNA RBM20 model of DCM while siRNA-MTSS1 model predicted appearance does not approach the improvement seen in siRNA TTN or siRNA CSRP3, in model of DCM. Note: For all graphs p-values with dotted lines represent a pairwise Student’s t-test (two sided). Shere multiple doses were tested, dashed lines with rightward arrow represent a linear model for a dose-dependent effect of siRNA MTSS1 upon model probability.

The primary clinical deficit in DCM is decreased cardiac contractility. In order to further explore the *in silico* observations and hypotheses generated from high throughput imaging we examined the effect of MTSS1 knockdown in a variety of iPSC-CM models of DCM using a commercially available engineered heart tissue (EHT) system. EHTs seeded with cells carrying a known pathogenic mutation in TTN P22353X display impaired contractility relative to isogenic controls [Fig.S4] and when TTN P22353X EHTs were treated with siRNA for MTSS1 (siMTSS1) we observed an improvement of 129% in twitch force (p=1.9e-04) [Fig.3A] and additional experiments confirmed improvements in twitch force were correlated with the degree of siRNA mediated reduction in MTSS1 levels [Fig.S5]. Next, we surveyed the impact of siRNA-MTSS1 in other forms of genetic DCM. Improved contractility was observed in iPSC-CMs with siRNA knockdown of CSRP3/MLP treated with siMTSS1 [Fig.3B] as well as with an isogenic line of RBM20 carrying a known pathogenic mutation S635FS^18^ where we observed a 284% improvement in twitch force (p<2e-16). Notably there was no improvement in sarcomere appearance in quantitative microscopy and no corresponding improvement in contractility in BAG3 iPSC-CM models of DCM treated with siRNA MTSS1 [Fig.S6]. The observation of improved twitch force confirmed the possibility that MTSS1 knockdown may have a beneficial effect upon cardiac contractility in DCM related to pathogenic mutations in TTN, CSRP3, and RBM20.

**Figure 3.**
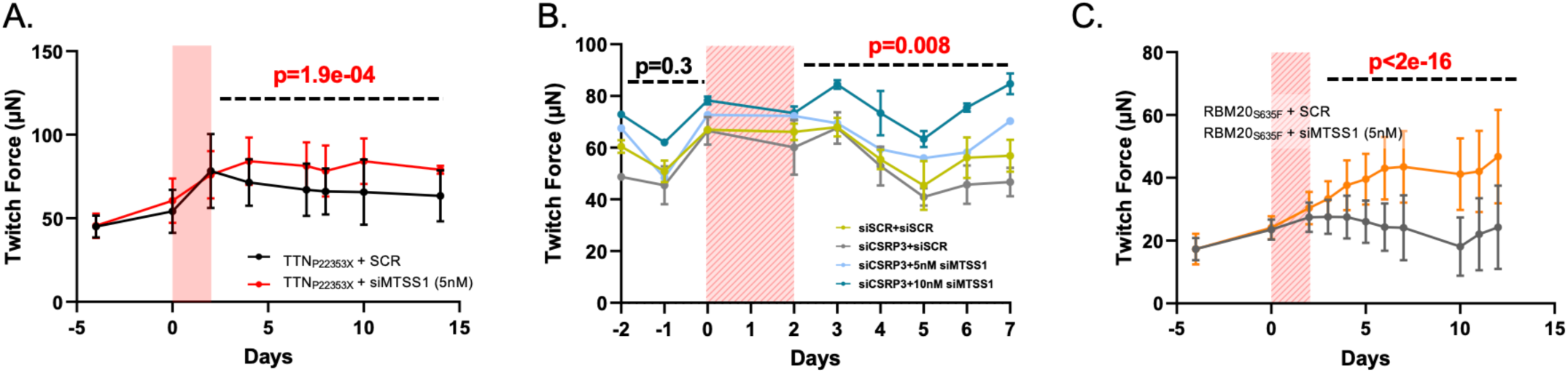
siRNA Knockdown of MTSS1 Improves Contractility in iPSC-CM EHT Models of DCM. **A.** Engineered Heart Tissues (EHTs) seeded with iPSC-CMs carrying the TTN_P22353X_ were treated with a mixture of three siRNAs knockdown of MTSS1 and show a sustained improvement in twitch force relative to the SCR control (p=0.003). In a repeat experiment individual MTSS1 siRNAs were tested and the amount of knockdown of MTSS1 correlates with the improvement in twitch force [Fig.S5] **B.** EHTs seeded with WT iPSC-CMs treated with one of four conditions (SCR control) show that co-treatment of siRNA CSRP3 (a Mendelian cardiomyopathy gene) with a single efficacious siRNA for MTSS1 shows improvement in contractility in a dose dependent manner (p=0.008). Note that in the pre-treatment there were no significant differences in twitch force between dosage groups (p=0.3). siRNA MTSS1 treatment of EHTs seeded with RBM20 iPSC-CM line carrying the S635FS mutation show dramatic improvement in contractility (p<2e-16). Dashed lines represent the period over which p-value was calculated for the treatment effect of siRNA MTSS1 in a linear model accounting for repeated measures over time from single wells. Red hatched area represents the two day period of treatment with siRNA.

### Quantitative Microscopy and Mechanistic Investigations

To gain a better understanding of the role of MTSS1 within the cardiomyocytes and sarcomere biology, we examined the quantitative microscopy data for all siRNA and genetic iPSC-CM backgrounds tested. Notably with siRNA knockdown of MTSS1 we observed a reproducible increase in the total number of sarcomeres and the number of sarcomeres per cardiomyocyte across different genetic backgrounds [Fig.4A-C, Fig.S7]. Given the observation that siRNA knockdown of MTSS1 appeared to increase the number of sarcomeres within an iPSC-CM and improved measures of contractility in select iPSC-CM models of DCM, we sought to further investigate the molecular mechanism within cardiomyocytes by which MTSS1 exerts effects on sarcomeres and contractility. Confocal imaging of GFP labelled MTSS1 in iPSC-CMs suggested a peri-nuclear cytoplasmic localization of MTSS1 away from the periphery of the cell without significant co-localization to the sarcomere [Fig.5A].

**Figure 4.**
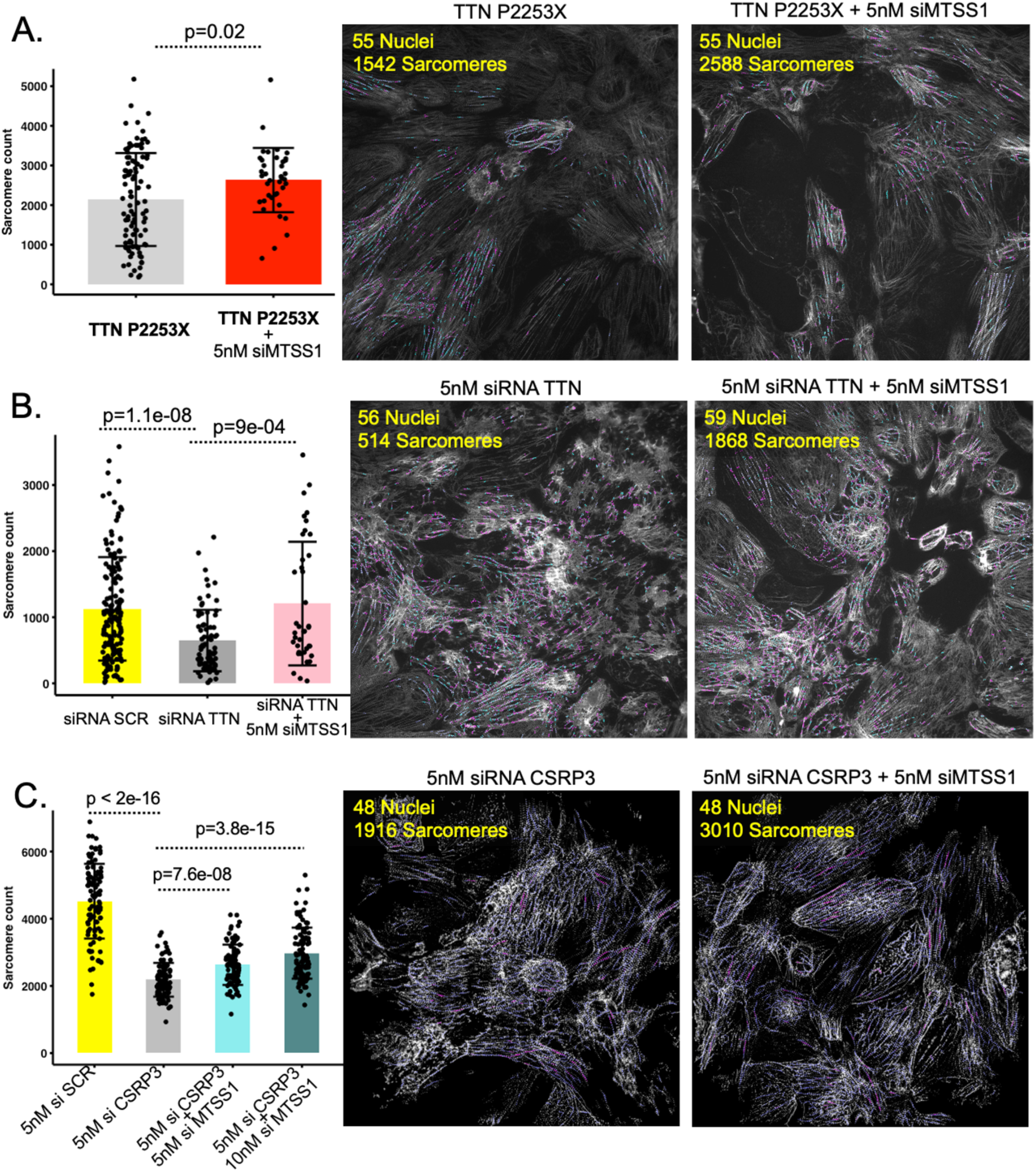
siRNA Knockdown of MTSS1 Increases Sarcomere Count in TTN and CSRP3 models of DCM. Example outputs from the quantification assay showing the identification of individual sarcomeres (colorized) derived from the ACTN2 and MYBPC3 staining of iPSC-CMs [Fig 2]. Nuclei and sarcomere counts per image are displayed in inset yellow text. Note that sarcomere counts of up to 3-5 fold difference may only appear as subtly different to the naked eye. **A.** An increase in sarcomeres in TTN_P22353X_ cell line is observed with 5mM siRNA MTSS1 treatment. **B.** In the siRNA TTN experimental condition, an increase in sarcomeres is observed with 5mM siRNA MTSS1 treatment with restoration of sarcomere levels to WT cells treated with a scrambled control. **C.** An increase in sarcomeres is observed to display a dose-dependent response to siRNA MTSS1 in the siRNA-CSRP3 experimental condition. For all graphs p-values with dotted lines represent pairwise Student’s t-test (two sided).

**Figure 5.**
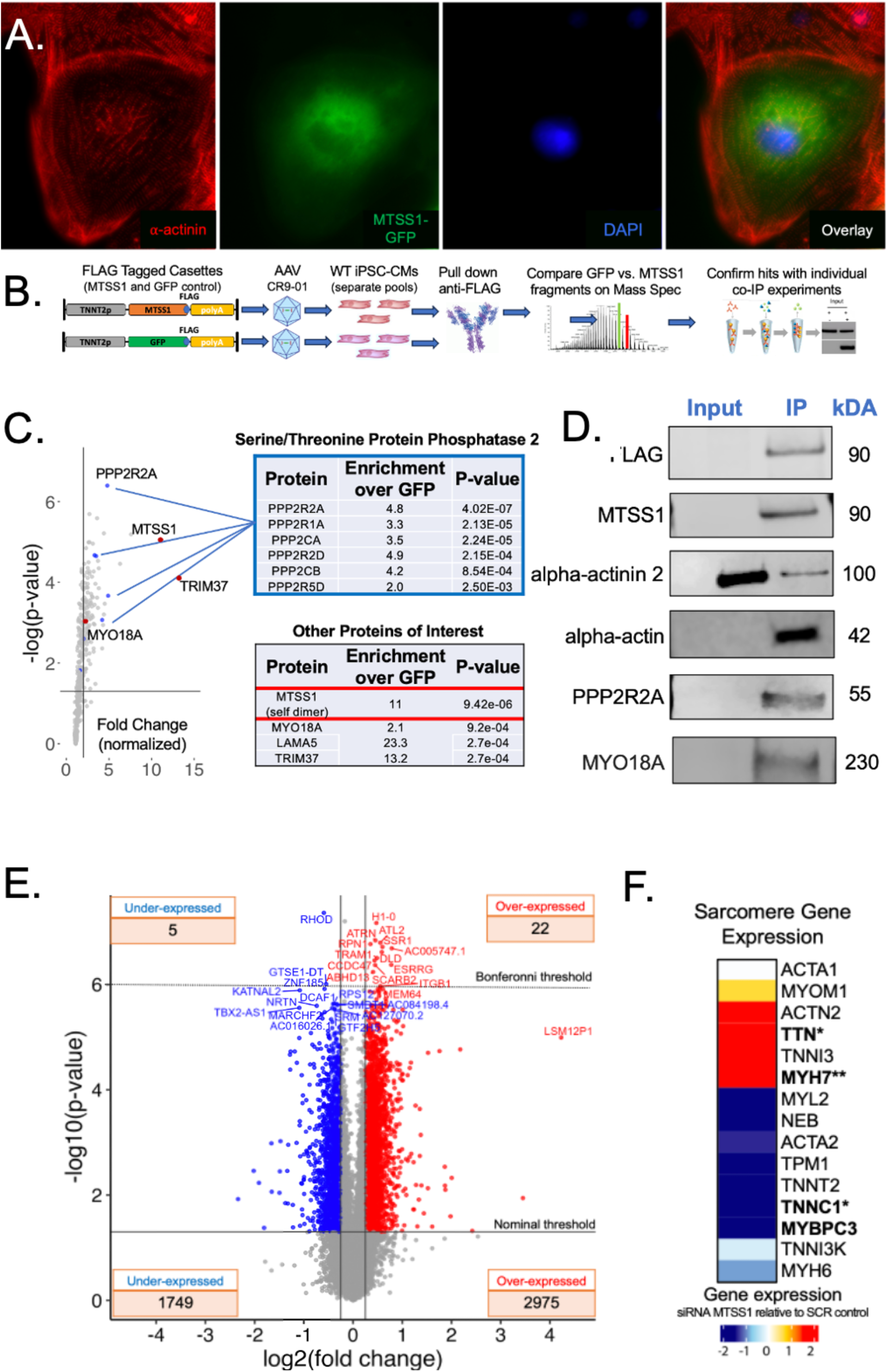
MTSS1 Displays Protein Interactions Indicative of Direct and Indirect Impact on Sarcomere Function and Transcriptional Response of MTSS1 Knockdown by siRNA is Consistent with a Primary Effect Upon Cardiomyocyte Contractility. **A.** Confocal imaging of GFP-tagged MTSS1 in iPSC-CMs Suggests a Peri-nuclear Cytoplasmic Subcellular Localization Without Significant Co-Localization to the Sarcomere. MTSS1 (labeled in green) is primarily visible in the cytoplasm, without localization to the cell membrane or nucleus (blue). GFP-labeled MTSS1 does not display significant co-localization with sarcomeres (overlay) that are clearly visualized by ACTN2 staining (red). Note that MTSS1 localization appears to be unevenly distributed throughout the cytoplasm, with more fluorescence observed in the center of the cell away from the periphery. **B.** Schematic of unbiased survey of MTSS1 protein-protein interactions. AAV are prepared with cassettes expressing MTSS1 or GFP and a FLAG tag under the control of the TNNT2 promoter and transfected into WT iPSC-CMs. After pull-down with an anti-FLAG antibody, mass spectrometry compares the relative abundance of digested protein fragments from each experiment, followed by confirmation of individual proteins with co-immunoprecipitation assays with protein-specific antibodies. **C.** Volcano plot of log_2_ fold change (normalized to sample and fragment abundance) and the -log p-value of enrichment in iPCS-CMs treated with AAV-MTSS1 relative to AAV-GFP. Inset boxes represent proteins of interest including members of the PPP2 signaling complex as well as MTSS1 self-dimer, LAMA5 and MYO18A. Purple colored dots indicate a significant number of ribosomal proteins. Data for all enriched proteins is available in the supplement. **D.** Individual co-IP experiments confirmed known and suspected interactions with ACTA and ACTN2, along with PPP2R2A and MYO18A. **E.** Volcano plot of the transcriptional response of TTN_P22353X_ EHTs treated with siRNA-MTSS1 uncovers key overexpressed genes related to hypertrophy SCARB2 and CCDC47 **F**. Heatmap of transcriptional response in select sarcomere genes in TTN_P22353X_ EHTs treated with siRNA-MTSS1 relative to untreated TTN_P22353X_ controls shows a robust increase in expression of TTN (p=4.9e-04, Q=0.01) and MYH7 (p=2.9e-06, Q=0.001) and decreases in expression of contractile inhibitory proteins TNNC1 and MYBPC3.

Formation of new sarcomeres in mature cardiomyocytes appears to arise from actin stress fibers within the periphery of cardiomyocytes^19^. Existing single cell data suggests that as a cytoskeletal-related actin binding protein MTSS1 is known to have relatively ubiquitous expression including cardiomyocytes [Fig.S9A] and other than actin is not previously known to have binding partners which modulate contractility or sarcomere function in cardiomyocytes [Fig.S9B]. To interrogate the protein binding partners of MTSS1 specific to cardiomyocytes, we performed AAV infection of a wild-type iPSC-CM cell line with a FLAG-tagged MTSS1 construct followed by a pull-down and mass spectrometry [Fig.5B]. Relative to an AAV transfection of FLAG-tagged GFP control, the MTSS1 construct appeared to interact with high-affinity to previously known binding partners^20,21^ [Table S1], multiple members of the protein phosphatase 2A complex, and notably the non-motor myosin MYO18A, a protein known to potentiate the formation of actin-stress fibers^22,23^ [Fig.5C]. To assess these findings, individual co-immunoprecipitation assays confirmed a protein-protein interaction in iPSC-CMs between MTSS1 and PPP2R2A and MYO18A alongside ACTN2 and ACTA2 suggesting a direct interaction between MTSS1 and the key components of actin-stress fibers in formation of sarcomeres [Fig.5D].

To better understand the performed RNA sequencing of EHTs seeded with TTN_P22353X_ iPSC-CMs comparing the transcriptional response of EHTs treated with siRNA-MTSS1 compared to siRNA scrambled control. Unbiased analyses uncovered upregulation of genes which are known to be involved in cardiac hypertrophy and calcium signalling such as TMEM64 and CCDC47^24–26^ [Fig.5E] as well as the mechanical response of cardiac fibroblasts to hypertrophy^27^. Importantly we found upregulation in contractile sarcomere proteins [Fig.5F] most notably siRNA MTSS1 mediated increases in TTN (0.41 log_2_FC, p=4.9e-04) and MYH7 (0.29 log_2_FC, p=2.9e-06), alongside decreases in inhibitory components of the sarcomere including TNNC1 the Calcium sensing subunit of the cardiac troponin complex (−0.16 log_2_FC, p=0.004) and MYBPC3 (−0.11 log_2_FC, p=0.01). Additionally, we observed upregulation in key Calcium handling, Desmosome, and Wnt/Hippo signalling genes and downregulation of apoptosis genes, as well as downregulation of heart failure marker NPPA (−2.5 log_2_FC, p=0.05) [Fig.S10]. The transcriptional response of increased sarcomere, calcium, and hypertrophy related genes to MTSS1 knockdown is consistent with the observation of a greater number of sarcomeres and increased twitch force.

## DISCUSSION

In a multi-modal set of analyses combining human genetics with human model systems of cardiac disease, we provide evidence supporting a relationship of lower cardiac MTSS1 levels to beneficial outcomes in three Mendelian forms of DCM. Our findings suggest the outlines of a Cardiomyocyte-specific mechanism by which MTSS1 is involved in regulating sarcomere production or turnover via interactions with actin stress fiber related protein MYO18A and ACTN2 and ACTA^22^. New sarcomere formation from actin stress fibers is a cytoplasmic process localized to the periphery of cardiomyocytes^19^, and we observed a peri-nuclear cytoplasmic localization of MTSS1 which may suggest a specific role in suppressing or modulating the assembly of mature sarcomeres that is de-repressed with siRNA knockdown. Earlier exploration of MTSS1 as a potential drug target did not show a clear beneficial effect in a mouse model of Tpm1 DCM^11^, which is consistent with our findings *in vitro* where a beneficial effect was only observed for specific genetic forms of DCM. in TTN, CSRP3, and RBM20 respectively but not TPM1. Importantly the *in vitro* findings of improved contractility in EHTs modeling TTN DCM observed with siRNA knockdown of MTSS1 are consistent with the novel observation of improved survival from cardiovascular death or heart transplant within individuals affected with TTN DCM who carry common alleles clearly demonstrated to reduce the cardiac expression of MTSS1^8^. Overall, these data support the potential for therapeutic benefits of reducing MTSS1 levels in select forms of DCM defined by a specific Mendelian genetic etiology, and may suggest a therapeutic mechanism of action whereby MTSS1 limits the number of sarcomeres in a cardiomyocyte.

Current treatment for Mendelian DCM falls within standard of care for heart failure including beta-blockade, afterload reduction, and SGLT2 inhibition but importantly does include therapies which address the underlying causes of reduced contractility^28^. The repeated observation in GWAS of improved contractility and heart failure outcomes with the T allele of rs12541595 which confers lower cardiac MTSS1 levels highlights the importance of using human genetics to uncover novel targets for heart failure. However, it is worth noting that in addition to supporting target selection, our results also suggest that Human genetics can support the first steps of clinical development enabling hypotheses about patient populations where a target may or may not be effective^29^. Therefore, an increased understanding of the genetic determinants and modifiers of disease and phenotypes in humans is likely to provide strong hypotheses for both target selection as well as matching the specific targets to select patient populations in order to reduce the time and costs of creating new therapies^30^.

Our approach employed machine learning to capture data from high content microscopy followed by quantitative analyses, which enabled the detection of changes in the cardiomyocyte and sarcomere—this exemplifies a robust and reproducible analytical approach for assessing the validity of potential targets. In addition to identifying MTSS1 as a target for knockdown to ameliorate DCM, this study contributes to the growing understanding of genetic targets for heart failure and paves the way for future therapeutic development using similar strategies^31^.

Our findings are not without limitations. While a potential mechanism whereby MTSS1 expression limits sarcomere turnover or maturation in Cardiomyocytes is suggested by the findings of increased sarcomeres and interactions with key proteins, a direct mechanistic link to these processes is not yet revealed by our data. In general iPSC-CMs have proven to be useful models of contractile phenotypes of cardiomyocytes, however even the most mature *in vitro* systems do not replicate the complexities of cardiovascular maturation, physiology, and multi-system pathophysiology of the adult Human heart^32^. The findings also suggest interactions of MTSS1 with the Serine/Threonine Protein Phosphatase 2A complex which has a variety of well described roles in regulating cardiac physiology^33^ – therefore it is possible that MTSS1 may modulate sarcomere structure and function in multiple ways. The findings relating an increased number of sarcomeres to increased contractility may suggest that sarcomere production and/or turnover could be relevant clinical and translational phenotypes for heart failure and cardiomyopathy in general. Further work on the relationship of MTSS1 to the sarcomere structure and cardiomyocyte function is necessary.

Additional limitations are inherent to the nature of the findings from human genetics; the observation of protective effects of lower MTSS1 in Human TTN DCM is enabled by a large number of individuals in the UK Biobank with pathogenic mutations in TTN, the most prevalent of Mendelian genetic form of DCM. While the observation of protective effect is striking and supported by the experimental data reported here, the protective allele for MTSS1 is present since conception and does not represent an intervention though the effect on DCM risk is assumed to be outweighed by the Mendelian TTN allele. This observation of a protective effect requires replication in another longitudinal dataset of individuals affected with TTN DCM, and unfortunately no evidence derived from human genetics is yet available for RBM20 or CSRP3 which are much less prevalent causes of monogenic DCM than TTN.

In summary, our findings illustrate the power of human genetics to select specific targets for cardiovascular disease as well as to identify patient populations affected with Mendelian DCM that are likely to benefit from manipulation of a specific targets. These data suggest that careful quantitative approaches to Cardiomyocyte biology can predict meaningful phenotypic responses of EHTs, which are in turn representative of observations from Human genetics. Together these data link Human genetics with *in vitro* Human model systems to illustrate the potential for quantitative approaches to cardiovascular discovery.

## METHODS

### Survival Analyses and Human Genetics

Genetic and clinical data in the UKB cohort were obtained from the UKB (https://www.ukbiobank.ac.uk). The UKB was approved by the North West Multi-Centre Research Ethics Committee and all participants provided written informed consent to participate in the study. DCM was defined as ICD-10 diagnosis code I42.0 or ICD-9 code 425.4. Statistical analysis was performed with R software using the *survival* package. Kaplan-Meier curves were produced with survfit function and hazard ratio (HR) was obtained from Cox regression model adjusted for sex and age at DCM diagnosis.

### hiPSC Culture and Maintenance

WT hiPSCs used in this study were derived from the WTC-11 line which were modified with a blasticidin resistance gene at the MYH6 locus. These cells were further modified with CRISPR to introduce a TTN P22353X^+/-^ mutation to create the DCM mutant line. hiPSCs were cultured in E8 medium (Life Technologies) on Matrigel (Corning) coated plates at 37°C and 5% CO_2_ and medium was changed daily until cells achieved a confluency of approximately 80%. Cells were then passaged using 0.5 mM EDTA in PBS (Invitrogen) as needed until differentiations.

### hiPSC Differentiation into hiPSC-CMs

hiPSCs were differentiated into ventricular cardiomyocytes using a Wnt-modulating protocol [Lian et al., 2012]. In brief, hiPSCs were first induced with 7 μM CHIR990921 (LC Labs) in RPMI+B27(−insulin) (Invitrogen). Two days post-induction, cells were placed in RPMI+B27(−insulin) for 24 hrs, followed by RPMI+B27(−insulin) supplemented with 5 μM IWP-2 (Sigma-Aldrich) for 48 hours. At 7 days post-induction, cells are then fed with RPMI+B27. At 9 days post-induction, cells are cultured in RPMI+B27 + 1 μM blasticidin (Gibco) for 9 days to select for differentiated cardiomyocytes. Purified cells were frozen down in CryoStor10 (StemCell Technologies) 19-21 days post-induction. Only cardiomyocytes that were >90% cTnT^+^ as verified by flow cytometry were used in this study.

### hiPSC-CM Cell Culture

hiPSC-CMs were thawed and added to a 384-well plate at a density of 10,000 cells/well. Wells were coated with 1:100 ratio of Matrigel up to 24 hours in advance. Edge wells were avoided to minimize edge-effects and cells were allowed to recover for 7 days with media changes every 2-3 days with RPMI+B27. After 7 days, cells were transfected with 5nM siRNA using Opti-MEM and Maturation Media. Media with the siRNA was removed after 2 days and media changes occurred every 2 to 3 days post siRNA treatment for 7 days. After 7 days, media was removed from the plate and wells fore fixed using 4% PFA. Wells were then stained for α-actinin (1:200 anti-ACTN2 Rabbit monoclonal Invitrogen 701914) and cMyBPc (1:200 anti-mybpc3 mouse monoclonal Sana Cruz sc137237) and Hoechst nuclear dye.

### Quantitative Microscopy Analyses

As described above, sarcomere proteins (α-actitin, and cMyBPc) are fluorescently tagged with AF+594 and AF+488(Alexa Flour, ThermoFisher) respectively allowing for high-throughput imaging of sarcomeres using an IXM confocal microscope (Molecular Devices San Jose CA). Five non-overlapping images were taken per well in a cross pattern avoiding edges and visualized using the FITC, TxRed, and DAPI channels. Images were read into a custom MATLAB script ‘Tamarack’. Learning from our previous sarcomere image analysis where we used a two-class deep learning model based on healthy and diseased iPSC-CMs (PMID), Tamarack is a modified program incorporating aspects of SarcTrack (ref: PMID 30700234), Tamarack employs three parallel Morlet wavelets with a set distance range, *d,* between peaks to represent the pattern of a sarcomere. The use of a three parallel wavelets reduced the incident of false positive sarcomeres compared to the traditional double wavelet pattern. Images were initially processed by masking for sarcomeres using either channel to remove unstained space and improve analysis time. Masked images are contrast corrected and binarized. The wavelet is then convolved over the image and rotated and re-sized to quantify sarcomere presence, orientation, and length. Tamarack reports sarcomere count, length, orientation, and pattern strength as well as variation of all these metrics. Sarcomere count is reported as both an absolute count as well as normalized to nuclear count to account for variation in the number of cells per well. Outputs were averaged from each of the five images per well and reported as an average. Successful siRNA knockdown of both background genes and target gene (MTSS1) were confirmed in separate experiments.

### Analysis of Tamarack Outputs

#### Random Forest/Sarcomere ‘scoring’ methods

Analysis of Tamarack Outputs were completed in R environment for statistical computing using the ‘randomForest’ package version 4.7-1 (citation DOI:10.1023/A:1010933404324). Raw data was cleaned by excluding conditions that resulted in “0” counts as well as the top and bottom 1% of all measured parameters. To inspect the data, Quantile Quantile plots were used to visualize the distribution of data and to determine if exclusion of additional outliers was necessary, and boxplots were used to visualize the quantile distribution, outliers, and mean of individual conditions, and compare amongst treatment groups.

Data was normalized by plate and split into training and validation datasets in a (0.7 to 0.3) ratio across multiple plates using the untreated (WT/CDI) to siRNA-knockdown and/or isogenic background for each Mendelian cardiomyopathy gene tested (TTN, MLP/CSRP3, BAG3, RBM20, TPM1). The random forest was constructed and trained upon the training dataset and the ‘ntree’, ‘classwt’, and ‘nodesize’ parameters were manually optimized to maximize the negative predictive value of the final model to 95% or better. The best performing model was then applied to the data comprising the treatment groups of interest, with the output set to estimate the probability of each condition (untreated WT/CDI or siRNA/isogenic backround). To assess the impact of a particular treatment group, the untreated WT/CDI probabilities were analyzed with either a student t-test or standard linear model (lm function in R) and visualized with a beeswarm plot (ggplot2 v.3.4.2, ggbeeswarm 0.7.2). An example dataset along with an analysis and visualization script are included in the online supplement along with the Tenaya Therapeutics Cardiovascular Genetics GitHub page [**TBD**].

#### Engineered Heart Tissues

Engineered heart tissues (EHTs) were fabricated using the commercially available Mantarray kit (Curi Bio) and manufacturer’s protocols were used. In brief, casting wells arrayed in a 24-well plate format were pre-filled with 50 μL of a 6.4 U/mL thrombin solution, and the post lattice was inserted into these wells. hiPSC-CMs and human cardiac fibroblasts (HCFs) were then mixed with 5 mg/mL of fibrinogen at a per tissue composition of 5×10^5^ cardiomyocytes and 7.5×10^4^ HCFs. This cell and fibrinogen solution was then added to the casting wells containing thrombin and thoroughly mixed around the inserted posts. Tissues were allowed to crosslink for 80-90 minutes at 37°C before being lifted out of the casting wells and placed into a new 24-well plate filled with fresh RPMI+B27 media. EHTs typically began beating within one week of casting. In experiments in which EHTs were transfected with siRNA, transfection occurred 18 days after tissue fabrication.

#### Confocal Microscopy

CDI cells were thawed and added to a 6 plate at a density of 1M cells/well. Wells were coated with 1:100 ratio of matrigel up to 24 hours in advance. After 96 hours, cells were transfected with 2.5 ug MTSS1-GFP construct using ViaFect (Promega E4981) reagent according to manufactural protocol. Media with the Viafect and DNA was removed after 24 hours and media changes occurred every 2 days post treatment for 4 days. After 4 days, media was removed from the plate and wells were fixed using 4% PFA. Wells were then stained for α-actinin (1:200 anti-ACTN2 Rabbit monoclonal Invitrogen 701914) and Hoechst nuclear dye and imaged on

## Data Availability

All data produced in the present study are available upon reasonable request to the authors

---

**Figure S1:**
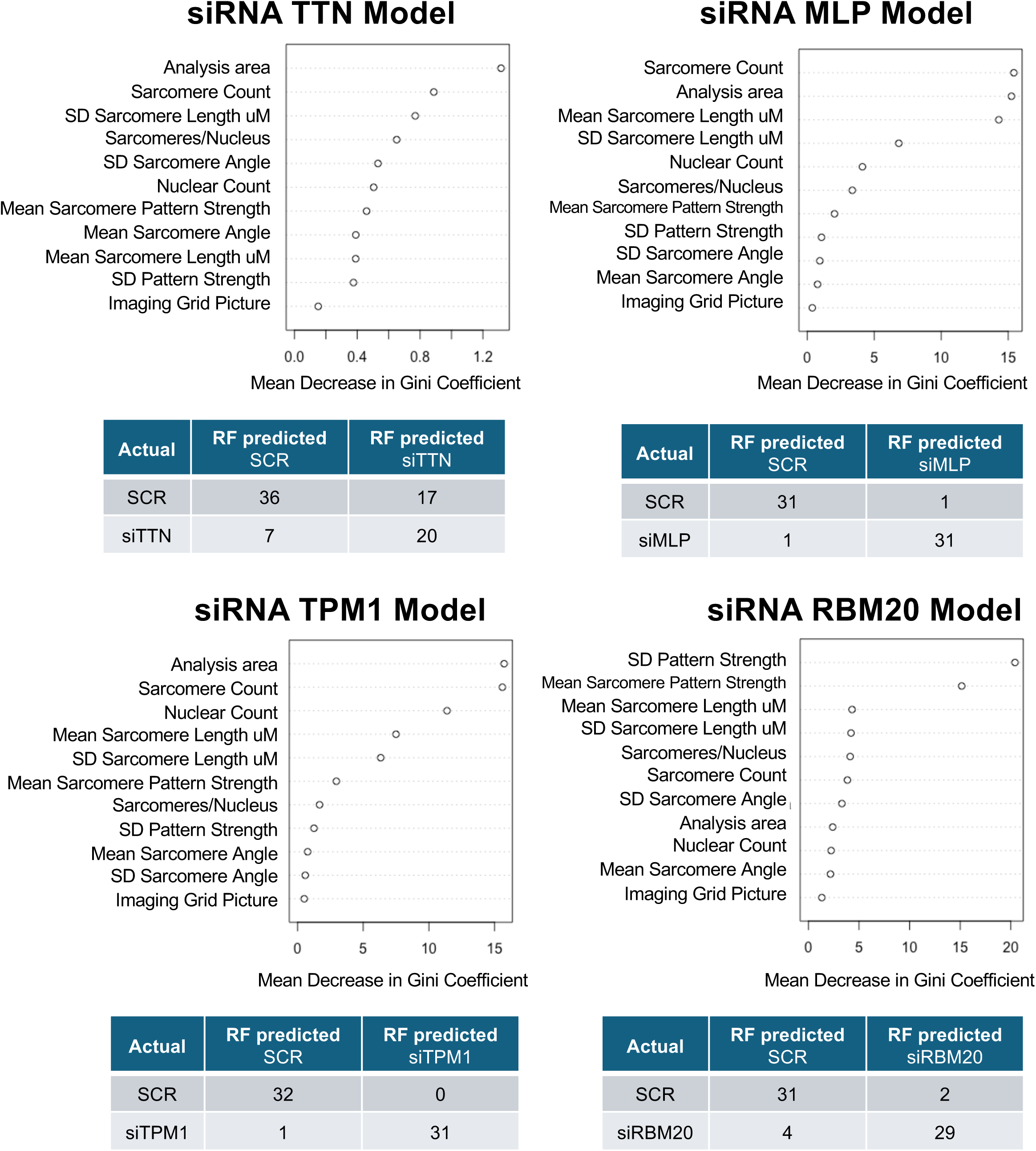
Diagnostic Information and Plots for Random Forest Prediction Models. For each Random Forest prediction model a variable importance plot is provided detailing the relative value of each predictor in the classification performance as measured by the Gini coefficient (inequality among the values of a frequency distribution between the siRNA-DCM and SCR groups). A confusion matrix of the model performance on the held-out test data is also presented to show model performance.

**Figure S2:**
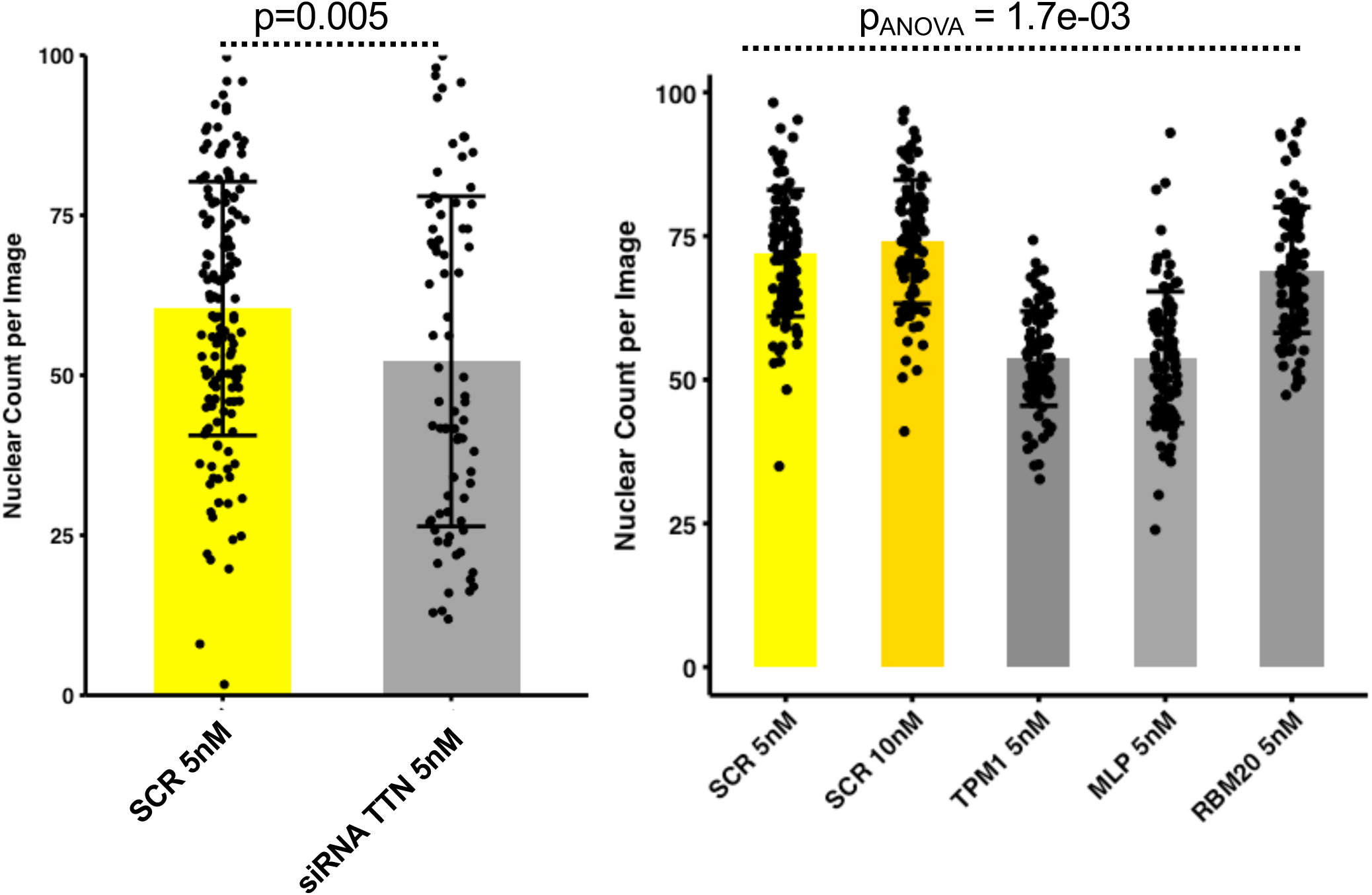
Treatment of WT iPSCs-CM with siRNA for DCM Genes May Cause Mild Toxicity. The number of nuclei per image are notably lower when treated with siRNA for different monogenic DCM genes. The siRNA-TTN experiment was performed separate from the other experimental conditions, therefore it is plotted separately. For all graphs p-values with dotted lines represent pairwise Student’s t-test (two sided).

**Figure S3:**
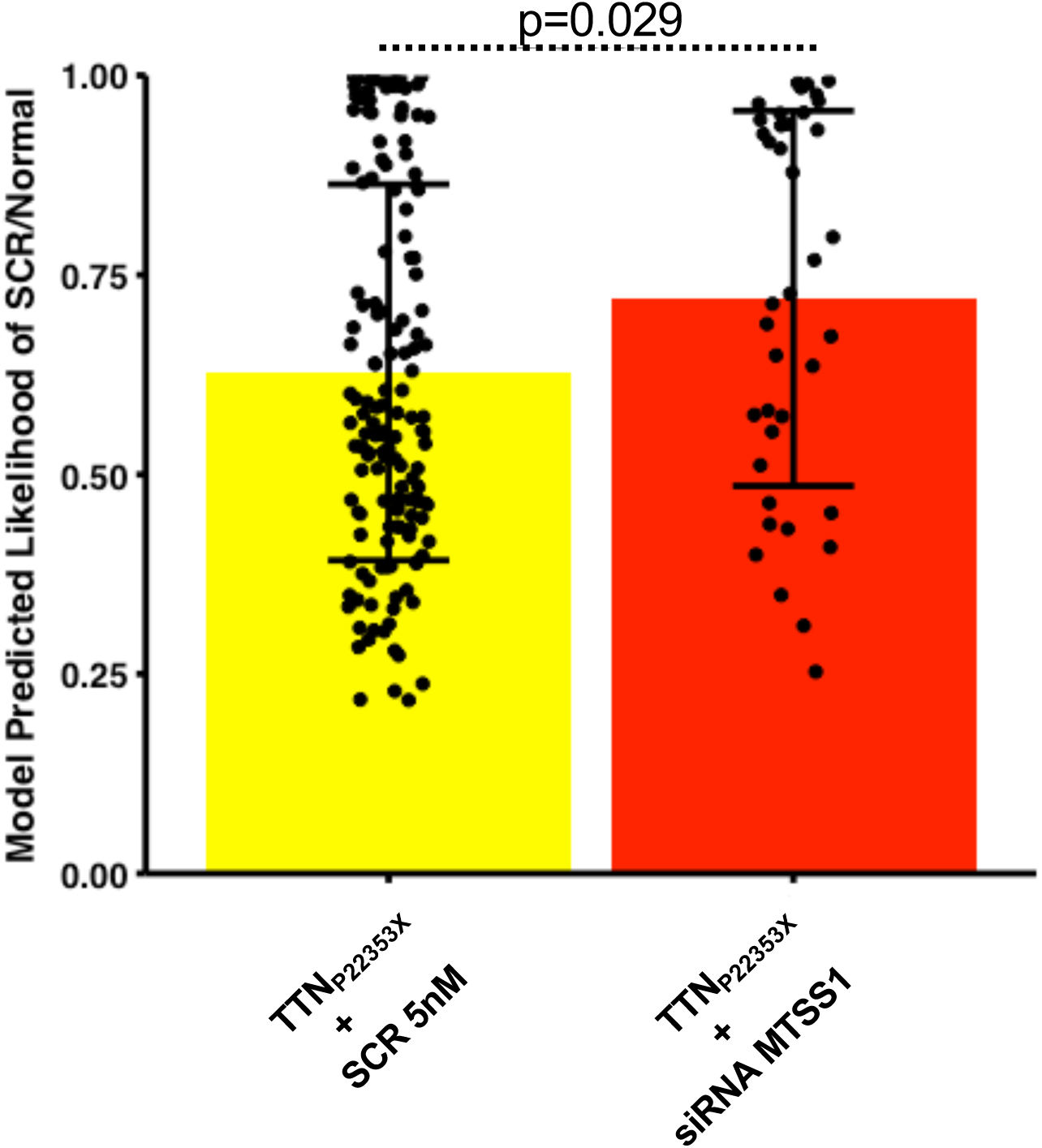
MTSS1 Knockdown by siRNA Shifts the Model Classification of iPSC-CMs Carrying the Pathogenic Mutation TTN_P22353X_ Causal for DCM. The random forest model from siTTN [Fig.S1] applied to high-content imaging from an isogenic TTN line treated with siRNA MTSS1 shows a shift in model classification away from disease and towards SCR/Normal in a statistically significant manner. For all graphs p-values with dotted lines represent pairwise Student’s t-test (two sided).

**Figure S4:**
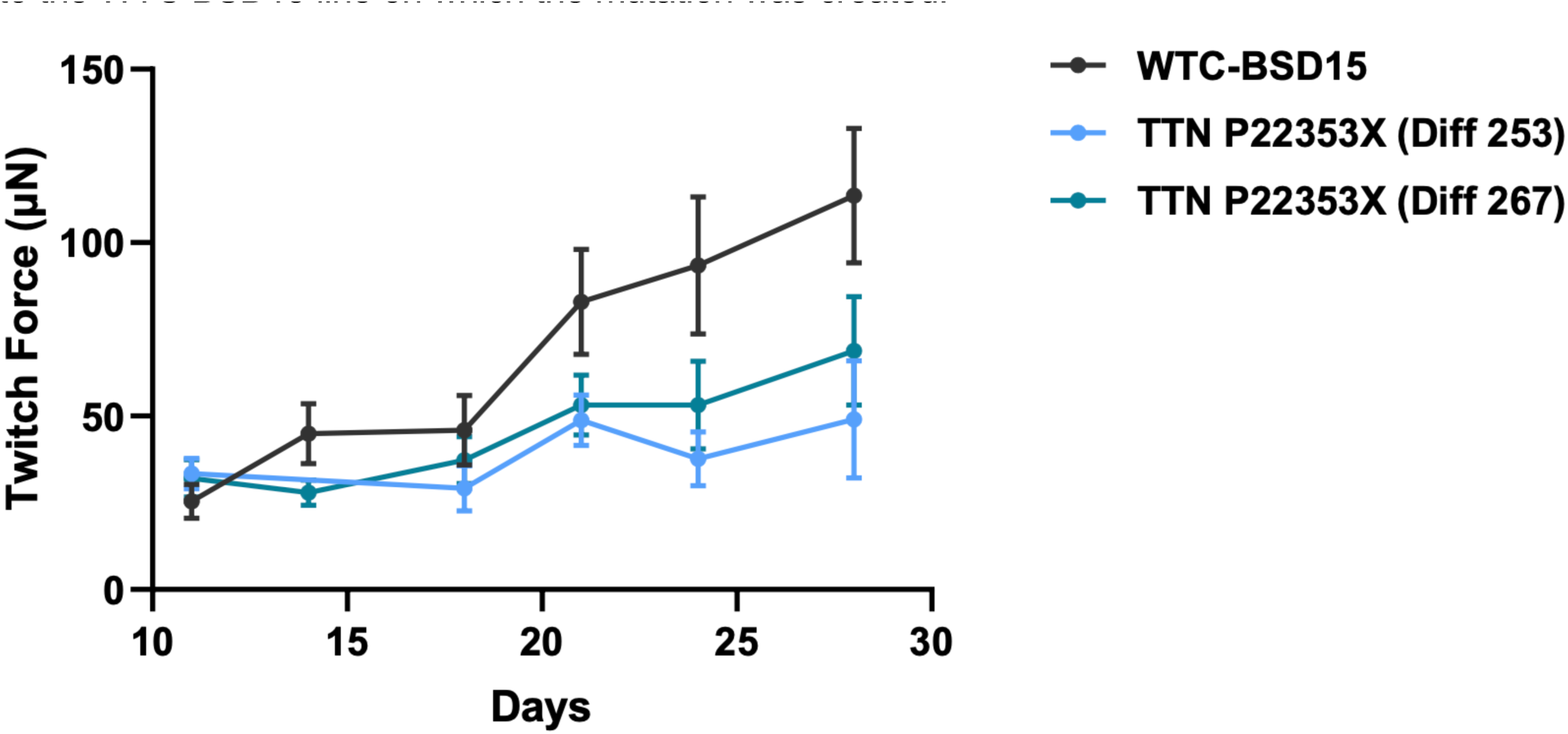
EHTs Seeded with iPSC-CMs Carrying the Pathogenic Mutation TTN_P22353X_ Causal for DCM Display Reduced Twitch Force Relative to Isogenic Controls. Two independent differentiations of the TTN_22353X_ show a sustained decrease in twitch force compared to the WTC BSD15 line on which the mutation was created.

**Figure S5:**
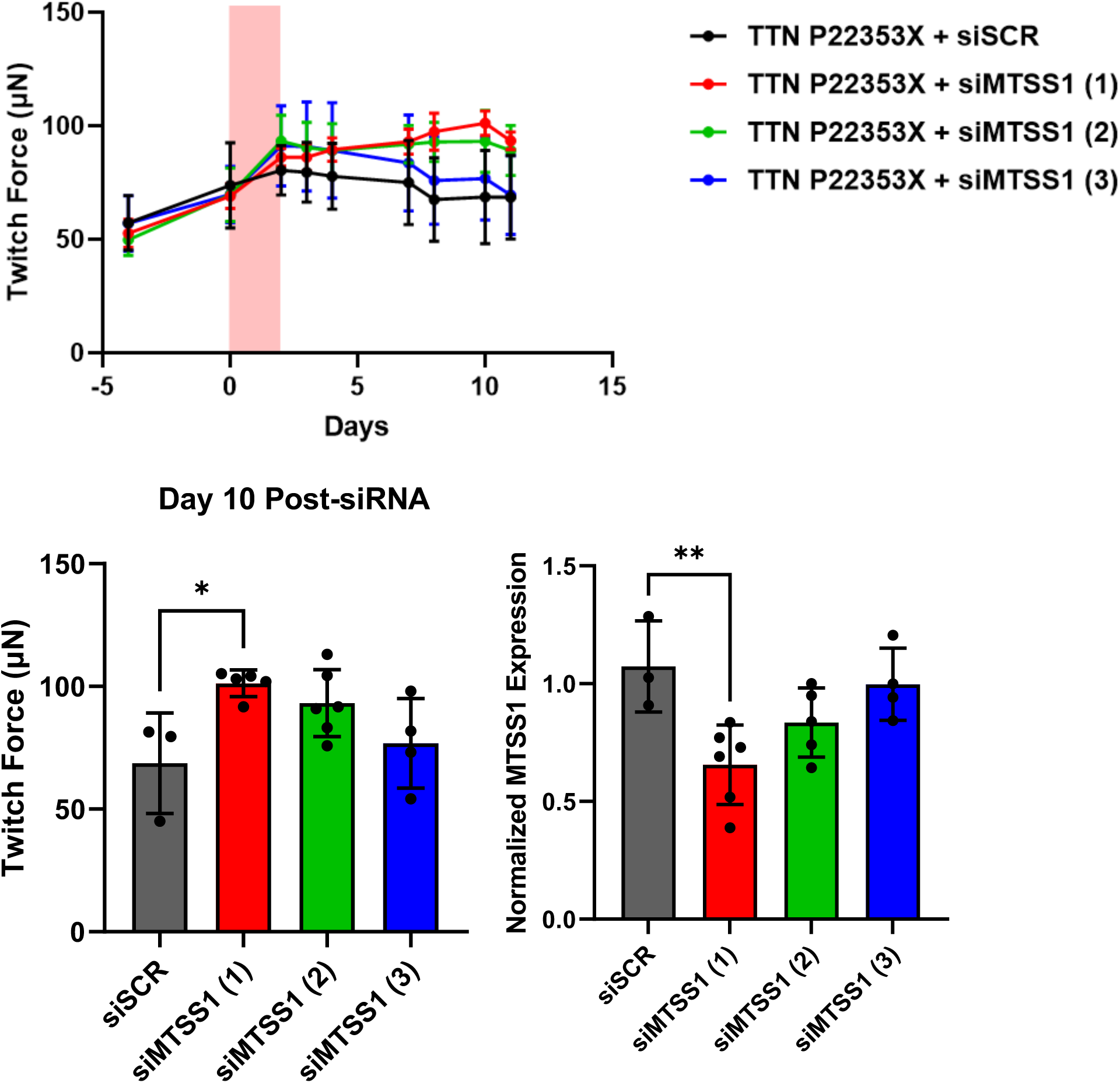
EHTs Seeded with iPSC-CMs Carrying the Pathogenic Mutation TTN_P22353X_ Causal for DCM Display Improvements in Twitch Force Relative to Efficacy of MTSS1 Knockdown of by Individual siRNAs

**Figure S6:**
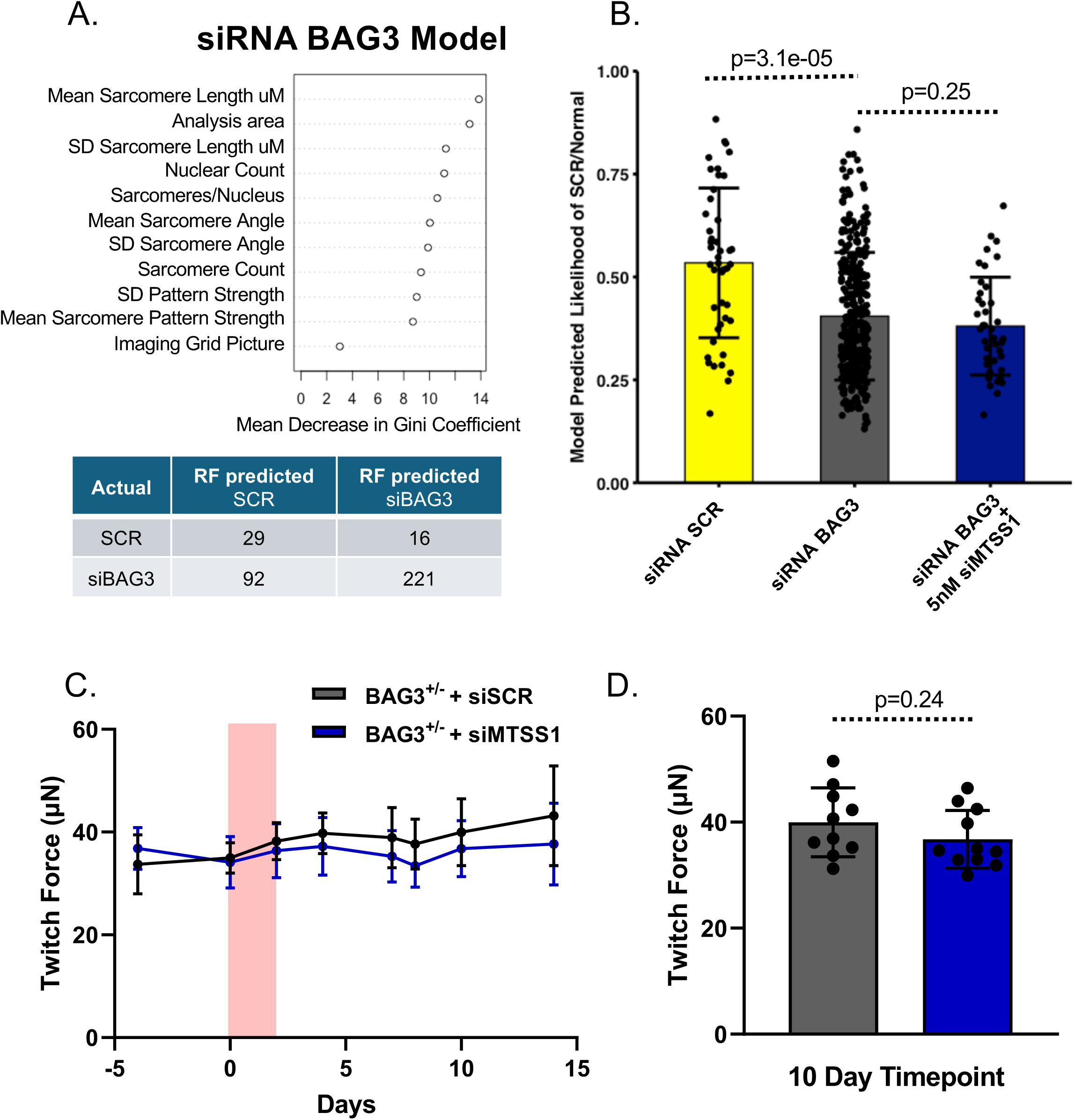
Knockdown of MTSS1 by siRNA Does Not Appear to Improve iPSC-CM Models of BAG3 DCM. A. Variable importance plot and confusion matrix of the siRNA BAG3 model performance on held-out test data. B. The random forest model from siRNA-BAG applied to high-content imaging from siRNA BAG3 cells shows adequate distinction between siRNA BAG and siRNA SCR control (p=3.1e-05) and no shift in model classification in siRNA-BAG3 treated with siRNA MTSS1 (p=0.25). C. Heterozygous BAG3 knockout EHTs treated with siRNA MTSS1 appear to show lower twitch force than untreated cells. D. A detail of the day 10 timepoint for EHTs in panel C. For all graphs p-values with dotted lines represent pairwise Student’s t-test (two sided).

**Figure S7:**
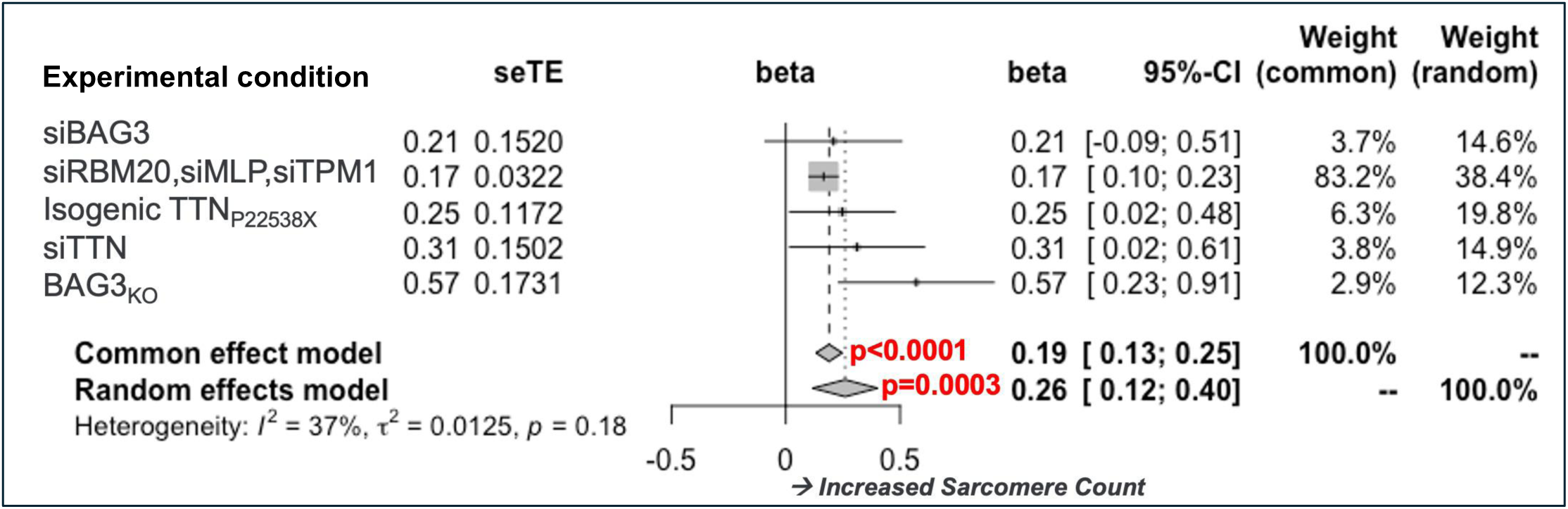
A Meta-Analysis of Quantitative Data Across Experimental Conditions Indicates that an Increased Number of Sarcomeres is a Shared Response of iPSC-CMs to siRNA Knockdown of MTSS1. For each of multiple experimental conditions and cell lines, sarcomere count was normalized to the condition and the effect of siRNA knockdown of MTSS1 analyzed. The standardized results from the analysis of each condition/cell line was included in a meta-analysis showing that across experimental conditions and cell types, knockdown of MTSS1 reproducibly and robustly resulted in an increase in the number of sarcomeres in both a common (p<0.0001) and random effects (p=0.0003) methods for meta-analysis.

**Figure S8:**
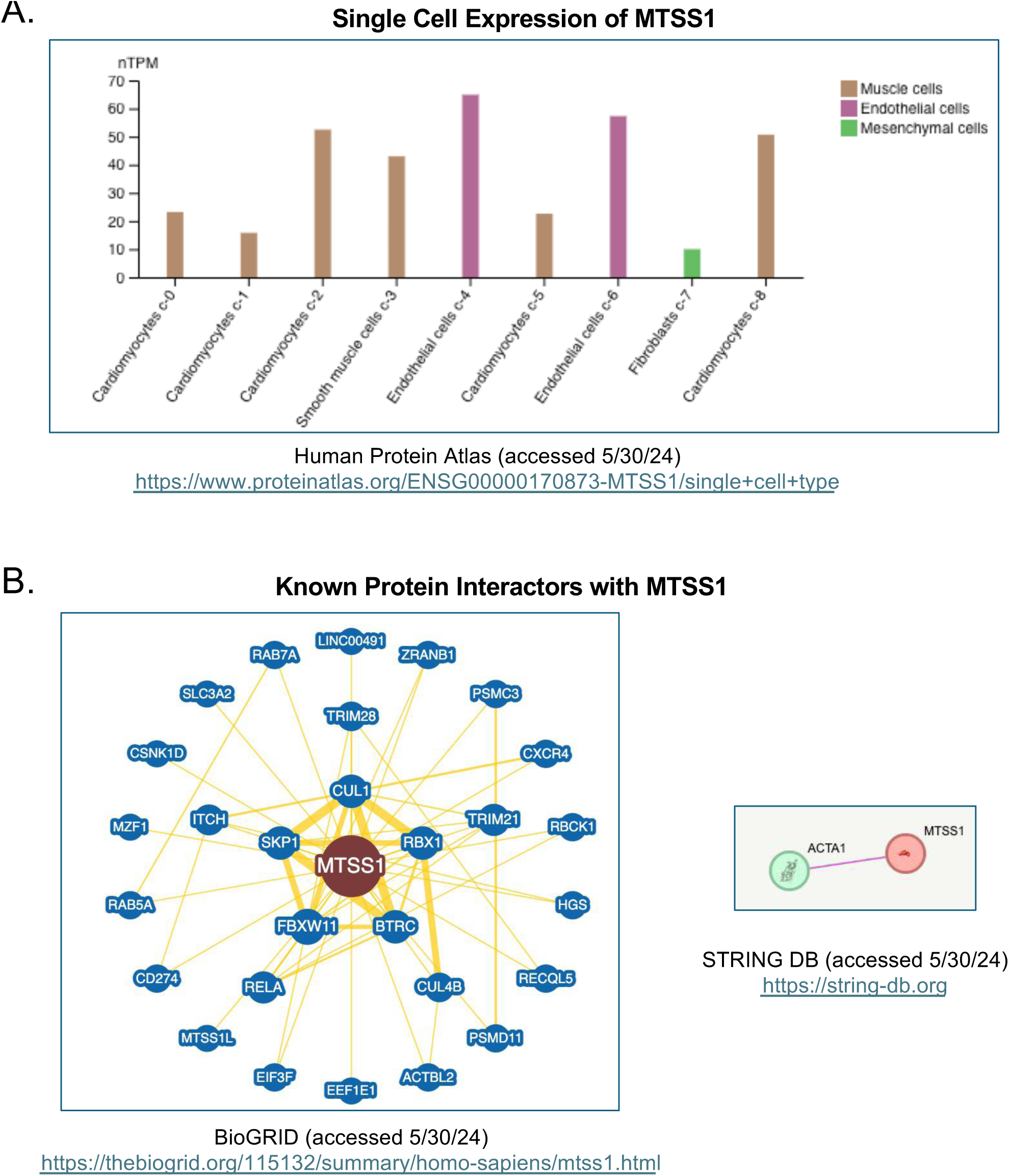
MTSS1 Is Expressed In Multiple Cell Types within the Heart and Does Not Have Known Binding Partners Related to Sarcomere Biology. **A.** Single cell expression of MTSS1 accessed from the human protein atlas shows moderate expression of MTSS1 in select groups of Cardiomyocytes and Endothelial cells within the heart and relatively low expression within Fibroblasts. **B.** Two protein-protein interaction maps (limited to experimental confirmation of physical interactions described in the scientific literature) identifies known interactors with MTSS1 from the STRING and BioGRID databases—no cardiomyocyte specific proteins are idenfied, though ACTA1 is known to have a role in both cardiomyocyte and skeletal myocyte sarcomere function.

**Figure S9:**
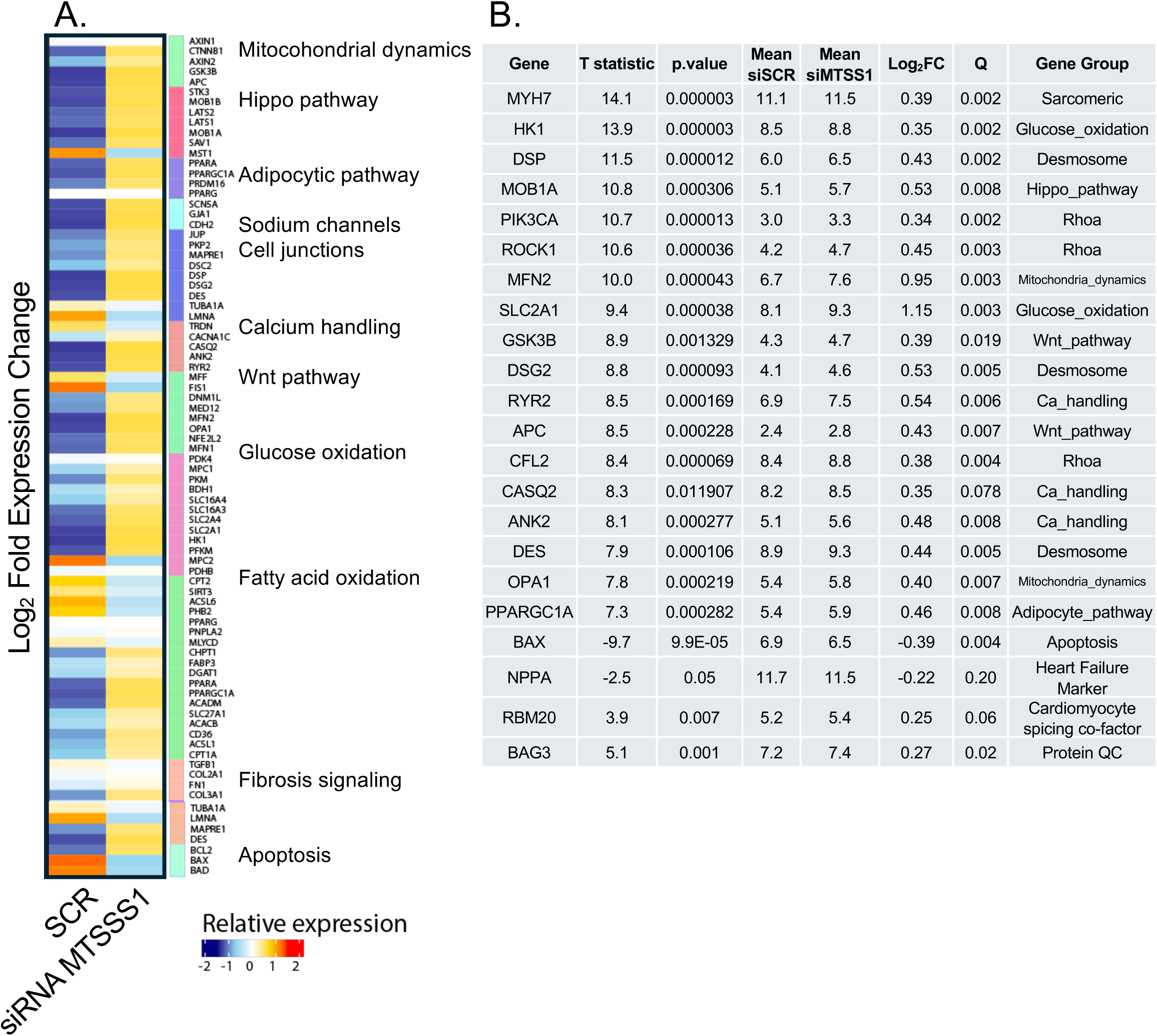
Transcriptional response of TTN_P22353X_ EHTs Encompasses Changes Across Pathways Related to Calcium Handling, Cellular Signalling, and Structure. A. Heatmap of SCR vs. siRNA-MTSS1 related transcriptional response organized by functional gene list. B. Select genes of interest representing key transcriptional drivers of pathways related to Calcium handling, Desmosome and Cell Junctions, and Wnt Hippo signalling. Of note heart failure marker NPPA is decreased in TTN_P22353X_ EHTs by siRNA MTSS1

